# Integrating Pulmonary and Systemic Transcriptomic Profiles to Characterize Lung Injury after Pediatric Hematopoietic Stem Cell Transplant

**DOI:** 10.1101/2025.03.31.25324969

**Authors:** Emma M. Pearce, Erica Evans, Madeline Y. Mayday, Gustavo Reyes, Miriam R. Simon, Jacob Blum, Hanna Kim, Jessica Mu, Peter J. Shaw, Courtney M. Rowan, Jeffrey J. Auletta, Paul L. Martin, Caitlin Hurley, Erin M. Kreml, Muna Qayed, Hisham Abdel-Azim, Amy K. Keating, Geoffrey D.E. Cuvelier, Janet R. Hume, James S. Killinger, Kamar Godder, Rabi Hanna, Christine N. Duncan, Troy C. Quigg, Paul Castillo, Nahal R. Lalefar, Julie C. Fitzgerald, Kris M. Mahadeo, Prakash Satwani, Theodore B. Moore, Benjamin Hanisch, Aly Abdel-Mageed, Dereck B. Davis, Michelle P. Hudspeth, Greg A. Yanik, Michael A. Pulsipher, Christopher C. Dvorak, Joseph L. DeRisi, Matt S. Zinter, the Pediatric Transplantation and Cell Therapy Consortium

## Abstract

Hematopoietic stem cell transplantation (HCT) is potentially curative for numerous malignant and non-malignant diseases but can lead to lung injury due to chemoradiation toxicity, infection, and immune dysregulation. Bronchoalveolar lavage (BAL) is the most commonly used procedure for diagnostic sampling of the lung but is invasive, cannot be performed in medically fragile patients, and is challenging to perform serially. We previously showed that BAL transcriptomes representing pulmonary inflammation and cellular injury can phenotype post-HCT lung injury and predict mortality outcomes. However, whether peripheral blood testing is a suitable minimally-invasive surrogate for pulmonary sampling in the HCT population remains unknown. To address this question, we compared 210 paired BAL and peripheral blood transcriptomes obtained from 166 pediatric HCT patients at 27 children’s hospitals. BAL and blood mRNA abundance showed minimal overall correlation at the level of individual genes, gene set enrichment scores, imputed cell fractions, and T- and B-cell receptor clonotypes. Instead, we identified significant site-specific transcriptional programs. In BAL, expression of innate and adaptive immune pathways was tightly co-regulated with expression of epithelial mesenchymal transition and hypoxia pathways, and these signatures were associated with mortality. In contrast, in blood, expression of endothelial injury, DNA repair, and cellular metabolism pathways was associated with mortality. Integration of paired BAL and blood transcriptomes dichotomized patients into two groups, of which one group showed twice the rate of hypoxia and significantly worse outcomes within 7 days of enrollment. These findings reveal a compartmentalized injury response, where BAL and peripheral blood transcriptomes provide distinct but complementary insights into local and systemic mechanisms of post-HCT lung injury.

## INTRODUCTION

Hematopoietic stem cell transplantation (HCT) combines high dose chemotherapy and/or radiation with intravenous infusion of hematopoietic progenitor cells in order to eradicate and replace malignant or dysfunctional cellular lineages.^1^ While the number of HCT procedures has increased dramatically and outcomes have improved over time, safety remains a concern. Specifically, acute or chronic lung injury can develop in 20-40% of patients due to chemotherapy toxicity, infections, and/or immune dysregulation.^2,3^ Post-HCT lung injury can lead to significantly impaired quality of life and premature death.^4,5^ Treatments largely include supportive care, anti-infectives, and attempts at immune modulation where possible, but are limited by a need for deeper understanding of the lung microenvironment after HCT.^6^

Therefore, the Pediatric Transplant and Cell Therapy Consortium (PTCTC) undertook a broad cross-sectional study of patients undergoing bronchoalveolar lavage (BAL) as diagnostic evaluation for post-HCT lung injury. We measured pulmonary microbiomes and paired human lung gene expression in BAL samples and identified four lung injury subtypes with varying degrees of dysbiosis, infection, inflammation, and cellular injury.^7^ Importantly, clinical outcomes varied significantly across the four subtypes, providing a biology-guided framework for risk stratification in this population. For example, patients with depleted pulmonary microbiomes, upregulated T-cell signaling, diminished alveolar macrophage activity, and signs of epithelial mesenchymal transition (EMT) had 3-4 fold higher in-hospital mortality than their counterparts.

It remains crucial to determine whether these processes are compartmentalized to the lung or are indicative of systemic disease, as this could affect the development of both precision medicine diagnostics and treatments. From a diagnostic standpoint, BAL is a medically invasive procedure that is limited to those with acceptably low illness severity and is not well-suited for serial sampling to monitor disease evolution or treatment-response.^8,9^ Thus, surrogates of lung biology obtained through noninvasive testing such as blood sampling could enable more nimble diagnosis and disease tracking. From a therapeutic standpoint, understanding whether and how these lung processes are reflected in or regulated by the blood compartment could inform whether eventual treatments ought to be delivered directly to the lung, through the blood, or both.

Thus, we undertook a systems biology approach to compare paired BAL and peripheral blood samples from pediatric patients with post-HCT lung injury. We questioned whether peripheral blood transcriptomes would correlate with key aspects of BAL transcriptomes that we had previously linked to disease evolution and clinical outcomes. Here we present an integrated approach to understanding BAL, blood, and combined signatures of post-HCT lung injury that advance our understanding of local and systemic disease processes after pediatric HCT.

## RESULTS

### Patients

As previously described, the PTCTC SUP1601 study enrolled 229 pediatric HCT patients who developed post-HCT lung injury and underwent 278 clinically-indicated BAL procedures.^7^ Patients were enrolled across 32 children’s hospitals in the United States, Canada, and Australia between 2014-2022. For this study, paired peripheral blood was collected during BALs and 210 BAL-blood pairs in 166 patients from 27 hospitals were included for analysis (**Figure 1**). Patients varied broadly in age, sex, race, ethnicity and geography; transplant indication was most commonly hematologic malignancy followed by receipt of mostly bone marrow or peripheral blood allografts from a variety of donor types (**Table 1**). Post-HCT lung injury signs and symptoms included hypoxia, dyspnea, declining pulmonary function testing, and/or abnormal chest imaging and often developed in conjunction with other complications such as graft versus host disease (GVHD). BAL was performed a median 131 days post-HCT (IQR 37-376), at which point the cohort displayed a median absolute neutrophil count (ANC) of 3,042 cells/uL (IQR 1,620-5,511) and a median absolute lymphocyte count (ALC) of 482 cells/uL (IQR 184-1,159); approximately half of the cohort required supplemental oxygen at the time of sample collection. Based on BAL results, lung injury was classified as lower respiratory tract infection (95/210), non-pulmonary sepsis (4/210), or idiopathic pneumonia syndrome (111/210). After each patient’s most recent BAL, 85 of 166 patients required intensive care (51%), 48 required ≥7 days of mechanical ventilation (29%), and 32 patients died prior to hospital discharge (19%).

**Figure 1.**
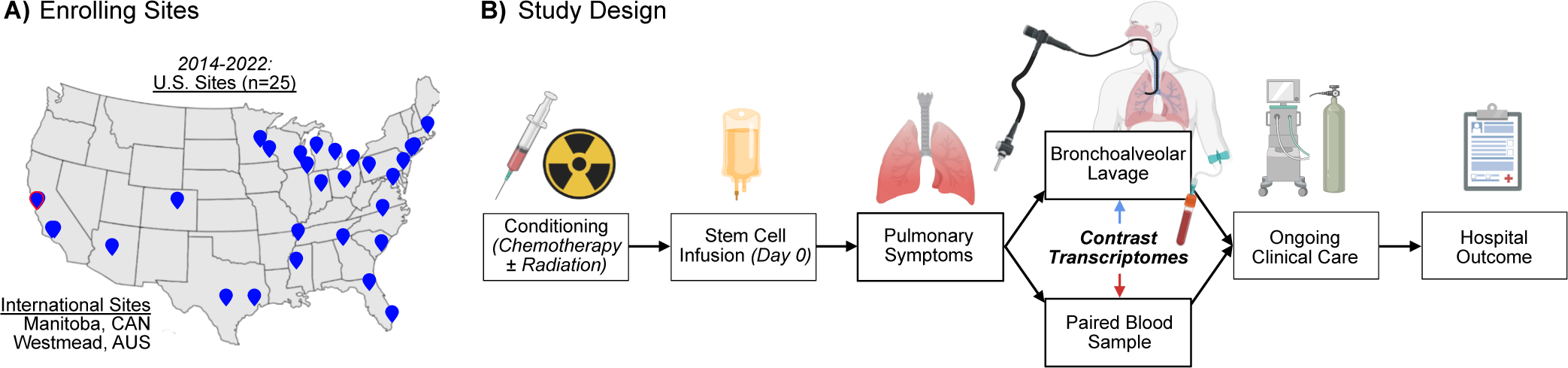
**(A)** Geographic location of participating children’s hospitals. **(B)** Patients were followed from the time of conditioning chemotherapy for the development of pulmonary symptoms. If bronchoscopy with bronchoalveolar lavage was planned for clinical reasons, patients were enrolled and BAL with a paired blood sample was collected. Patients were then followed clinically through hospital discharge.

### Contrasting Transcriptomes in Paired BAL and Peripheral Blood

We hypothesized that both BAL and peripheral blood could yield complementary insight into pulmonary disease states in this cohort. To contrast the information captured in each sample type, we first compared BAL and blood sample pairs. We observed tissue-specific differential gene expression, with 6,156 genes showing greater expression in BAL and 9,368 genes showing greater expression in blood (**Data File 1, Figure 2a**). For example, as expected, SFTPC was expressed to a greater level in BAL whereas HBA2 was expressed to a greater level in blood (**Figure 2b**). Many canonical lung-specific genes were detected in the blood of many samples (e.g. SFTPC 53/210, MUC5AC 92/210, LAMP3 180/210), albeit rarely above 1 read per million per sample.

**Figure 2.**
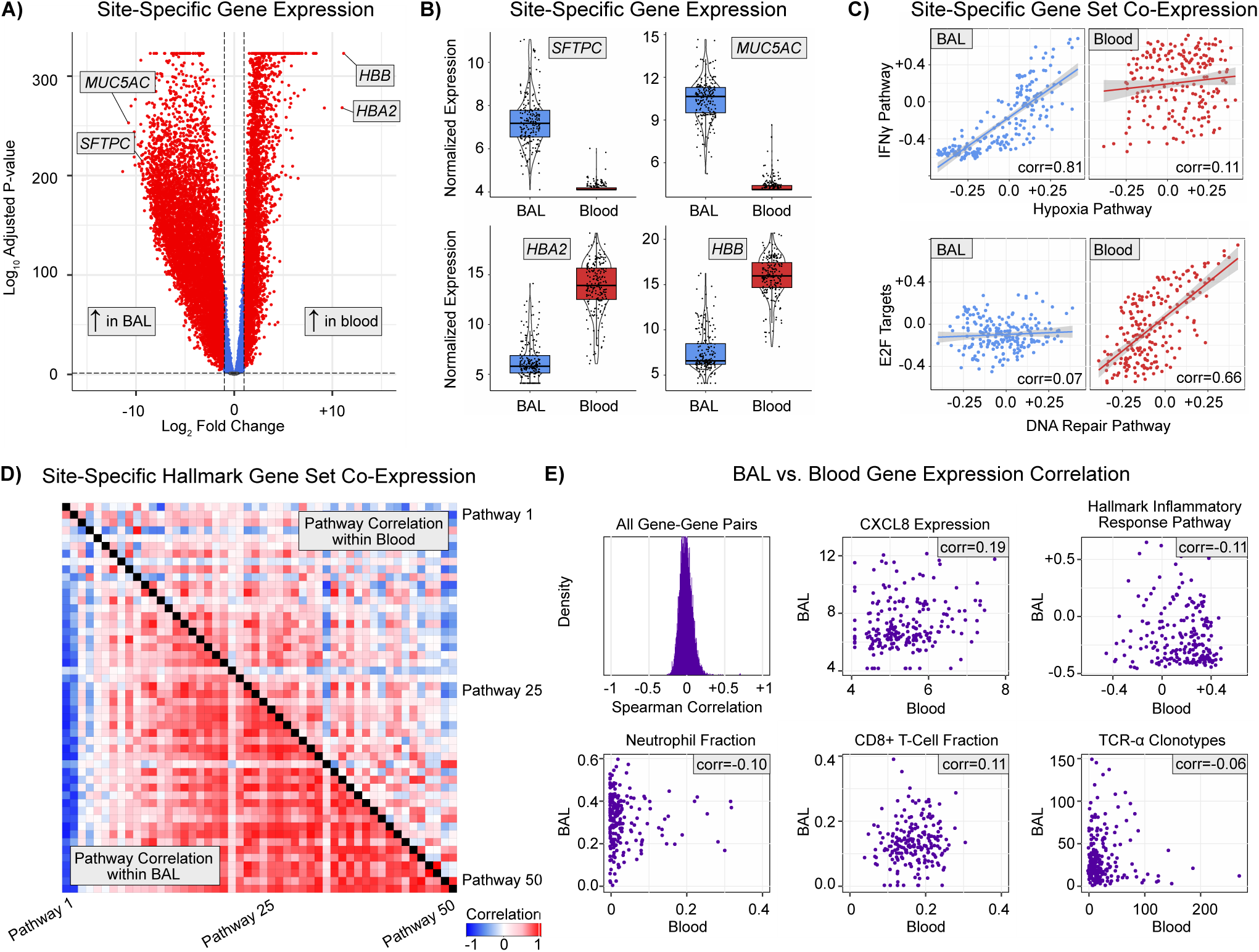
**(A)** Genes differentially expressed in BAL vs peripheral blood are shown. **(B)** Expression levels of select genes specific to lung (SFTPC, MUC5AC) and blood (HBA2, HBB). **(C)** Gene set enrichment scores to the 50 MSigDB Hallmark Pathways were calculated and correlation between expression of each gene set was calculated within each body site and then contrasted to identify site-specific coregulation. Here we show unique co-expression of Hallmark Hypoxia and IFNg gene sets in BAL but not blood, and unique co-expression of Hallmark DNA Repair and E2F targets in blood but not BAL. **(D)** Correlation of MSigDB Hallmark pathways within blood (top right triangle) and within BAL (bottom left triangle) are shown. See Supplemental Figure for detailed labels. **(E)** BAL-blood correlation was calculated for expression levels of n=7,169 protein-coding genes and the distribution of correlation coefficients is plotted. Expression levels of CXCL8 in BAL and blood are shown as an example of minimal correlation. Gene set enrichment scores for the MSigDB Hallmark Inflammatory Response gene set measured in BAL and blood are also shown as an example of minimal BAL-blood correlation. BAL and peripheral blood cell type fractions were imputed using CIBERSORTx and reference atlases, and cell fractions across body sites were correlated using Spearman correlation coefficients, with neutrophils and CD8+ T-cells shown as examples. T- and B-cell receptor clonotypes were measured using Imrep and the number of unique clonotypes across body sites were correlated using Spearman correlation coefficients, with TRA shown as an example.

To understand the different regulatory networks unique to each body site, we next tested for tissue-specific differential gene co-expression. When assessing enrichment scores to the 50 MSigDB Hallmark gene sets,^10^ differential co-expression networks revealed highly site-specific interactions (**Data File 2**). Of note, 39 gene sets were co-expressed in BAL but not blood; for example, Hallmark Hypoxia expression scores were correlated with Hallmark Interferon Alpha and Gamma signaling, DNA Repair signaling, and Oxidative Phosphorylation signaling in the lung (correlation ≥0.5) but not in the blood (correlation <0.1, FDR-adjusted p<0.05). In contrast, 9 gene sets were co-expressed in blood but not BAL; for example, Hallmark DNA Repair expression scores were correlated with E2F signaling in the blood (correlation ≥0.5) but not in the lung (correlation <0.1, FDR-adjusted p<0.05, examples in **Figure 2c**). Thus, overall, BAL gene networks showed much stronger co-expression than blood gene networks (**Figure 2d**), suggesting synchronization of biological pathways in the lung.

Given the site-specific differences in expression levels and co-expression networks, we next sought to determine the degree of cross-body-site transcriptome correlation. For any given gene, there was minimal correlation between BAL and blood expression levels (median Spearman rho -0.005, IQR -0.056 to -0.002, **Data File 3, Figure 2e**). As an example, the lack of BAL-blood correlation for CXCL8 gene expression is depicted in **Figure 2e**. Similar results were found when analyzing enrichment scores to broader gene networks in the MSigDB collection (**Data File 4,** example in **Figure 2e**) and when analyzing imputed cell fractions (**Data File 5, Figure 2e)**. For example, the fraction of neutrophils in BAL and blood sample pairs showed no correlation (Spearman correlation -0.099, FDR-adjusted p=0.520). In addition, the blood ANC correlated with imputed blood neutrophil fraction (Spearman rho=0.462, p<0.001) but not BAL neutrophil fraction (Spearman rho=0.043, p=0.549). We then calculated T- and B-cell receptor clonotypes using ImRep; we detected a median 16 TRA clonotypes in BAL (IQR 8-31, range 0-271) and 25 TRA clonotypes in blood (IQR 13-42, range 0-148) but no TRA clonotypes were shared across BAL-blood pairs (except 1 clonotype in one of 210 sample pairs). Similar results were found for TRB, TRG, TRD, and IGH, IGK, and IGL (**Data File 6**). Together these data highlight the uniqueness of the biological information contained in both BAL and blood.

### BAL and Peripheral Blood Transcriptomes Differ by Survival Outcome

To better understand site-specific and cross-site contributors to survival, we next contrasted transcriptomes in survivors and non-survivors. We again observed tissue-specific differential gene expression in non-survivors. In BAL, expression levels of 350 genes were associated with death; overall, non-survivors had increased expression of genes related to epithelial injury and hypoxia (e.g. SPP1, SFTPB, CEACAM6) and lower expression of genes related to innate immune signaling (e.g. CSF3R, CXCR1, IL1B, CXCL8, **Data File 7, Figure 3a**). In blood, expression levels of 656 genes were associated with death; non-survivors had increased expression of genes related to endothelial cell junction and smooth muscle activity (e.g. JCAD, PTGFR) and lower expression of genes related to lymphocyte signaling (e.g. JCHAIN, CCR7, **Data File 8**, **Figure 3b)**. Genes associated with death in BAL and in blood showed minimal overlap (**Figure 3c**), again indicating different site-specific contributors to outcome. Notably, CEACAM6, LGMN, and BCAT1 showed increased expression in both BAL and blood of non-survivors, while CD74, MUC5B, HLA.DRB1 and HLA.DPB1, and CFD showed lower expression in both BAL and blood of non-survivors.

**Figure 3.**
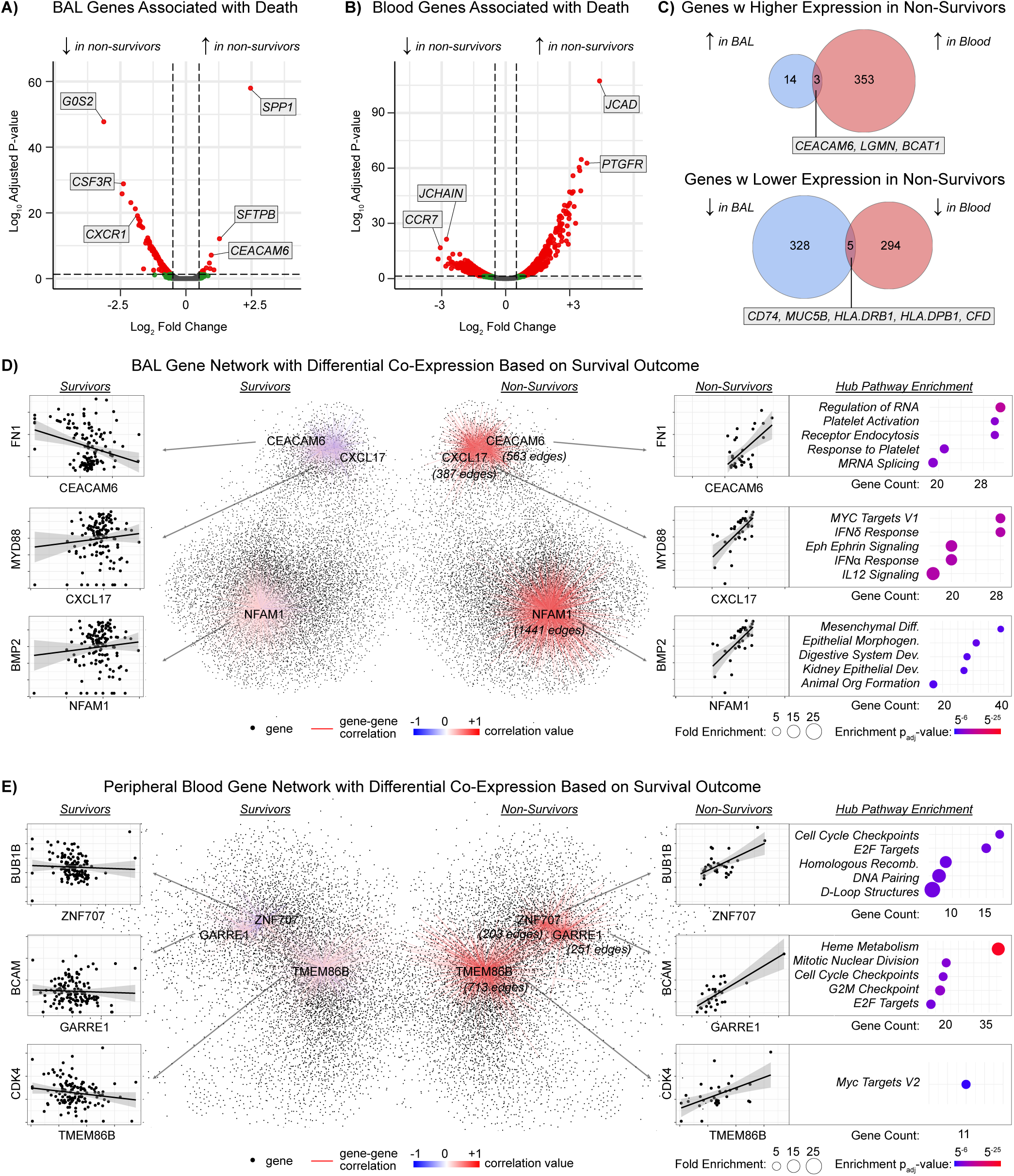
**(A)** BAL gene expression differences in non-survivors. **(B)** Peripheral blood gene expression differences in non-survivors. **(C)** Overlap between BAL and blood genes associated with mortality. **(D)** Network of genes co-expressed in BAL of non-survivors (right) but not co-expressed in survivors (left). Examples of hubs genes (CEACAM6, CXCL17, NFAM1) are shown. Examples of differentially co-expressed genes linked to each hub gene are shown (e.g. CEACAM6-FN1 co-expression) to illustrate differential gene-expression. To the right, pathway enrichment for hub genes are shown. **(E)** Network of genes co-expressed in peripheral blood of non-survivors (right) but not co-expressed in survivors (left). Examples of hub genes (ZNF707, GARRE1, TMEM86B) are shown. Examples of differentially co-expressed genes linked to each hub gene are shown (e.g. ZNF707-BUB1B co-expression) to illustrate differential gene-expression. To the right, pathway enrichment for hub genes are shown.

In many diseases, genes maintain stable expression levels but exhibit altered co-expression patterns with other genes, suggesting that they influence pathobiology through network reorganization rather than heightened expression.^11,12^ Therefore, we next tested for tissue-specific differential gene co-expression unique to non-survivors. In BAL, a network of 10,038 genes was co-expressed in non-survivors but these genes were not co-expressed in survivors (rho≥0.5 vs rho<0.1, FDR<0.05, **Data File 9**). Top hub genes included *CEACAM6*, which was co-expressed with 563 other genes related to platelet signaling and endoplasmic reticulum stress; *CXCL17*, which was co-expressed with 387 other genes related to immune signaling and hypoxia; and *NFAM1*, which was co-expressed with 1,441 genes related to EMT and lung morphogenesis (**Figure 3d)**. In contrast, in blood, a network of 6,929 genes was correlated in blood of non-survivors but not survivors **(Data File 10).** Top hub genes included *ZNF707*, which was co-expressed with 203 other genes representing cell cycle transitions and DNA repair, and *GARRE1*, which was co-expressed with 251 other genes representing heme metabolism (**Figure 3e**). These gene coregulatory networks complement the gene expression pathways noted above and accentuate the differences in biologic activity identified in BAL vs. peripheral blood specimens.

Although we observed minimal BAL-blood correlation in the overall cohort, we hypothesized that non-survivors might show greater cross-site correlation due to systemic illness and increased signaling across the alveolar-capillary membrane. However, we did not identify any gene sets that were differentially correlated across BAL and blood according to survivor status (all FDR>0.1, **Data File 11**). For example, Hallmark Hypoxia gene expression signaling in BAL and blood were minimally correlated for both survivors and non-survivors (p>0.05) and these correlations were not significantly different (FDR-adjusted p=0.994). This indicates that even the sickest patients do not show cross-site transcriptome correlation.

### BAL and Peripheral Blood Transcriptomes Differ by Lung Injury Subtype

We previously identified 4 subtypes of HCT-related lung injury based on BAL transcriptome-microbiome signatures.^7,13,14^ These subtypes varied not only in clinical outcomes, but also in burden of infection, microbiome dysbiosis, inflammation, and cell injury (**Figure 4a**). Of note, lung injury Subtype 1 was defined by alveolar macrophage (AM) predominance, a replete and balanced microbiome, minimal signs of inflammation, and superior clinical outcomes. To better understand the biology of these lung injury subtypes, we next contrasted BAL and blood transcriptome pairs in reference to the best-performing Subtype 1, with results summarized in **Figure 4 and Table 2** and described in detail below.

**Figure 4.**
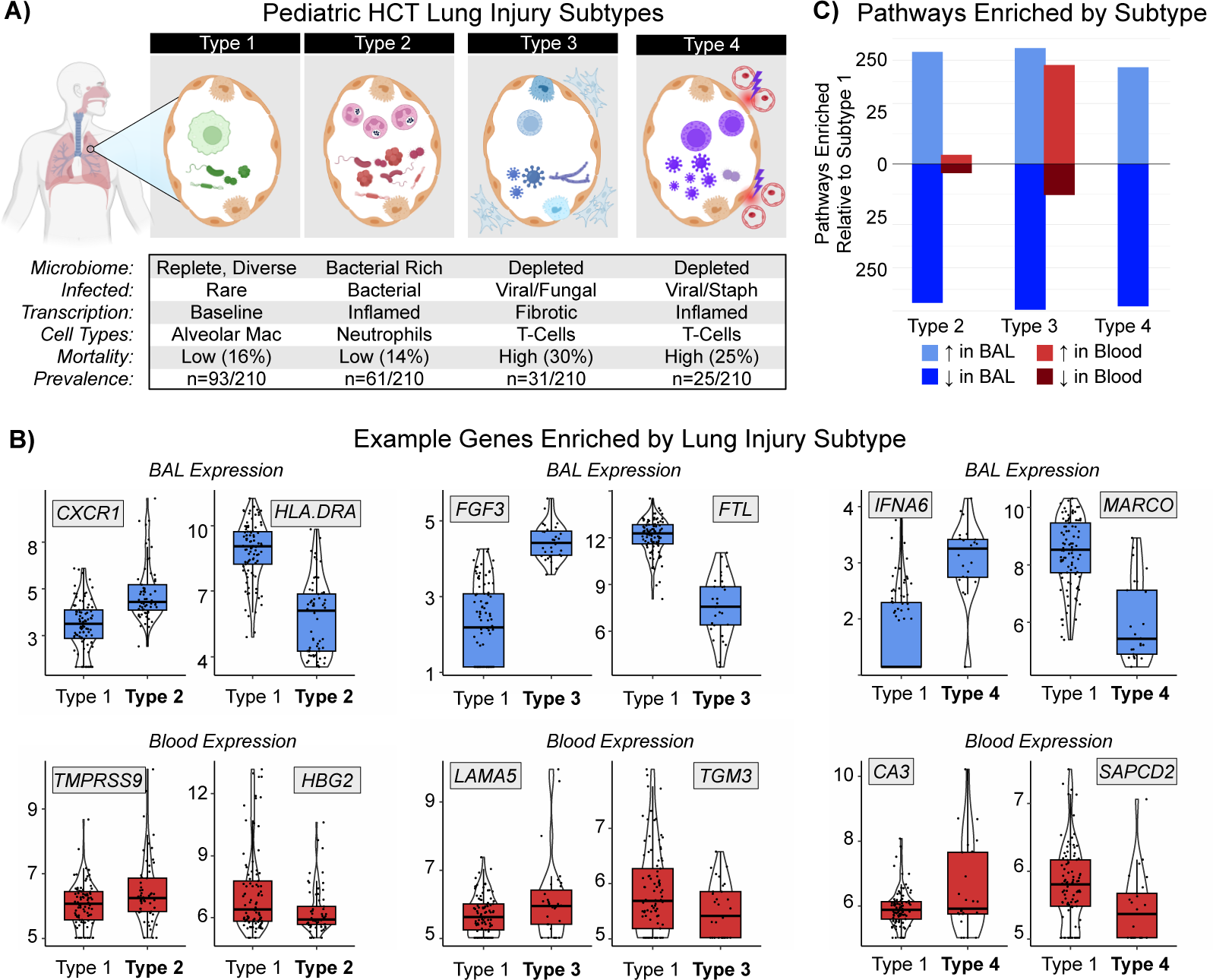
**(A)** Concept diagram for four post-HCT lung injury subtypes derived in the PTCTC SUP1601 cohort and validated in the University of Utrecht, the Netherlands cohort (*Nature Med 2024*). **(B)** Example BAL and blood genes differentially expressed in lung injury subtypes 2, 3, and 4 relative to subtype 1. **(C)** Differentially expressed BAL and blood genes underwent pathway analysis and pathways identified in BAL and blood gene are quantified to show greater overall differences detected in BAL as opposed to paired blood.

#### Subtype 2

We previously showed that patients with Subtype 2 had higher rate of pulmonary bacterial infections and low in-hospital mortality. ***In BAL:*** Subtype 2 patients differentially expressed 2,027 genes (|LFC|≥1), including higher expression of genes related to granulocyte and inflammasome pathways (e.g. CXCR1, IL1R2, S100A8) and lower expression of genes related to macrophage and lymphocyte signaling (e.g. CCL18, APOE, HLA.DRA, **Figure 4b**). Supporting this, cell deconvolution showed an increase in BAL neutrophil fraction and a decrease in the AM fraction relative to Subtype 1 (**Data File 12**). Patients with Subtype 2 had unique BAL co-expression of a network of 9,181 genes including top hubs NLRP3, CXCR1, and ALPL, which together showed enrichment for immune activation, cell-cell adhesion, platelet activity, and extracellular matrix interactions (**Data File 13**). ***In Peripheral Blood:*** Subtype 2 patients differentially expressed 1,061 genes, including higher expression of genes related to thyroid activity (e.g. CALCA, TPO, DIO2) and lower expression of genes related to heme metabolism (e.g. HBD, HBG2) with no differences in blood cell fractions noted (**Data File 14**). These patients also had greater blood co-expression of a network of 8,347 genes including top hubs NUP210, CDH13, and COL6A6, which together represent DNA replication, oxidative phosphorylation and cellular metabolism (**Data File 15**). Comparison of enriched pathways in BAL and blood showed only 1 overlapping gene set: *Hallmark Heme Signaling* was lower in both BAL and blood of patients with Subtype 2. These data illustrate that in patients with Subtype 2, pulmonary bacterial infections and a pulmonary neutrophil response are associated with a systemic neurohormonal response marked by increased blood cell metabolic activation without clear shifting of the fractions of blood cell types.

#### Subtype 3

In contrast, patients with Subtype 3 showed a high rate of pulmonary microbiome depletion as well as high mortality. ***In BAL:*** Subtype 3 patients differentially expressed 8,166 genes, including higher expression of genes related to fibroblast activation, myogenesis, and nitric oxide signaling (e.g. FGF3, BMP1, NOS) and lower expression of genes related to macrophage signaling (e.g. CD74, FTL, HLA.DRA) and response to hypoxia (e.g. HIF1A, HMOX1, **Figure 4c**). In conjunction, cell deconvolution showed an increase in BAL CD4 and CD8 fractions and a decrease in the AM fraction relative to Subtype 1 (**Data File 16**). Patients with Subtype 3 had unique BAL co-expression of a network of 6,724 genes including top hubs SIGLEC1, PTPRC, and HADHB, which together showed enrichment for collagen deposition, integrin interactions, and EMT (**Data File 17**). ***In Peripheral Blood:*** Subtype 3 patients differentially expressed 574 genes, including higher expression of genes related to collagen deposition and EMT (e.g. COL4A1, LAMA5, TIMP3), with no differences in blood cell fractions noted (**Data File 18**). These patients also had greater blood co-expression of 9,269 genes including top hubs ANO6, GPD2, and TLR8, which together represented DNA repair, T-cell signaling, and heme metabolism (**Data File 19**). Unique to Subtype 3, comparison of enriched pathways in BAL and blood showed significant overlap for genes related to EMT and endothelial activation. These data illustrate that in patients with Subtype 3, pulmonary microbiome depletion, T-cell activation, and pro-fibrotic signaling co-occur with systemic signs of EMT, suggesting a component of cross-body site communication regulating fibrosis pathways.

#### Subtype 4

Finally, patients with Subtype 4 showed both commensal microbiome depletion and viral infection, again with high mortality rates. ***In BAL:*** Subtype 4 patients differentially expressed 6,252 genes, including higher expression of genes related to natural killer/T-cell activity (e.g. IL2, KLRF1, IFNG, IFNA6, **Figure 4d**), beta-defensins (DEFB114, DEFB110), and EMT (COL11A1, MMP27), and lower expression of genes related to AM signaling (e.g. MARCO, FTH1, HLA.C), neutrophil signaling (e.g. MYD88, TREM2), and airway and alveolar epithelial function (e.g. SPRR3, MUC5B, SFTPB). In conjunction, cell deconvolution showed an increase in BAL CD4 and CD8 fractions and a decrease in the AM fraction relative to Subtype 1 (**Data File 20**). Patients with Subtype 4 had unique BAL co-expression of a network of 10,223 genes including top hubs SLC38A6, ETFDH, and TAF2, which together showed enrichment for collagen deposition, ankyrin interactions, and EMT (**Data File 21**). ***In Peripheral Blood:*** Subtype 4 patients differentially expressed only 156 genes, including weak overlap with IFNα/β pathways (e.g. OAS3, IFIT1) and modest increase in blood CD4+ fraction (**Data File 22**). These patients also had greater blood co-expression of 6,908 genes including top hubs PRR16, SCML2, and TRPC6, which together represented DNA repair, oxidative phosphorylation, and aerobic respiration (**Data File 23**). Comparison of enriched pathways in BAL and blood showed no overlapping gene sets. These data illustrate that in patients with Subtype 4, pulmonary microbiome depletion, T-cell activation, and epithelial injury response co-occur with non-specific systemic signs of increased metabolic activity, but BAL and blood transcriptomes overall showed minimal overlap.

#### Integrated BAL and Blood Transcriptome Signatures

Given that paired BAL and blood transcriptomes were not correlated, we next questioned whether they might would complement each other in understanding patient illness and clinical outcomes. By inputting normalized BAL and blood transcriptomes into multi-omics factor analysis (*MOFA)* followed by dimensionality reduction (*umap*), we detected two clusters of patients (**Figure 5a**). Cluster A consisted largely of patients from the lowest-risk lung injury subtype 1, whereas Cluster B consisted largely of patients from the sicker lung injury subtypes 2, 3, and 4 (**Figure 5b**). Patients in Cluster B were twice as likely to have required oxygen immediately prior to BAL (50.3% vs. 29.1%; 66/131 vs. 23/79, p=0.003) and when analyzing the most recent encounter for each patient, were twice as likely to be dead or require ongoing mechanical ventilation 7 days after BAL (37.0% vs. 17.2%; 40/108 vs. 10/58, p=0.008, **Figure 5c**). Consistent with the biologic themes identified by other methods above, Cluster B differentially expressed 9,784 genes in BAL (|LFC|≥1, FDR-adjusted p<0.05) representing alveolar macrophage depletion, lymphocyte activation, and epithelial injury, as well as 696 genes in blood, representing EMT and endothelial activation (**Data File 24**). Thus, this unified multi-omics approach demonstrates that post-HCT lung injury can be conceptualized as two large groups, with group B harboring mostly lung injury subtypes 2, 3, and 4 and showing greater illness severity and worse clinical outcomes.

**Figure 5.**
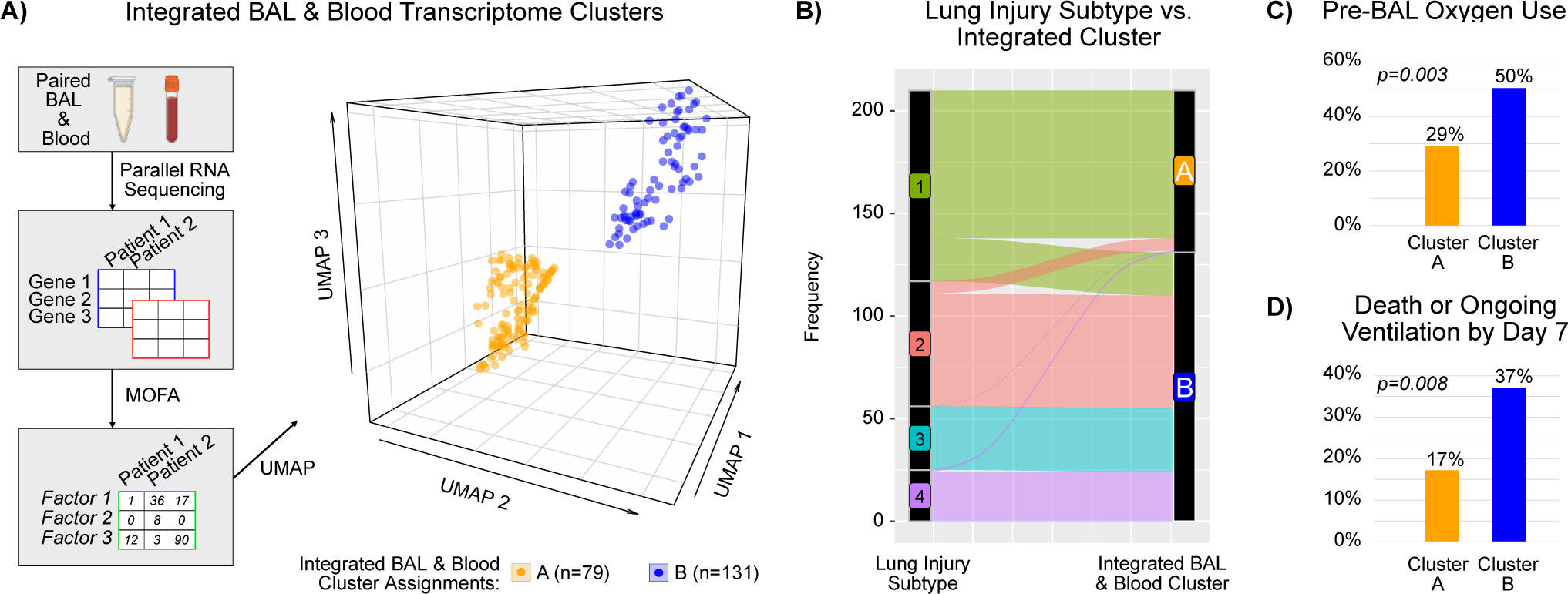
**(A)** Paired BAL and blood transcriptomes underwent multi-omics factor analysis (*MOFA*) followed by dimensionality reduction (*umap*) and k-means clustering to show 2 groups of patients. **(B)** Post-HCT lung injury subtype was mapped onto the two integrated transcriptome clusters, showing that most patients from subtypes 2, 3, and 4 mapped to Cluster B. **(C)** Approximately twice as many patients in Cluster B required oxygen prior to BAL sampling, and twice as many patients died or required ongoing mechanical ventilation within 7 days of BAL sampling.

## DISCUSSION

In this study, we compared 210 BAL and blood transcriptome pairs from pediatric HCT patients with lung injury. We identified minimal cross-site transcriptome correlation and instead uncovered unique site-specific transcriptome networks, suggesting body site compartmentalization of injury response. BAL signatures of mortality reflected immune perturbations, epithelial injury, and hypoxia signaling. Instead of mirroring the pulmonary compartment, peripheral blood transcriptomes instead showed broad signatures of endothelial injury, DNA repair, neurohormonal activation, and altered cellular metabolism, thus expanding our understanding of the systemic processes involved in post-HCT lung injury.

By contrasting different transcriptome programs in BAL and blood, we have learned about unique site-specific regulation. Generally speaking, in the BAL fluid, immune signaling was more tightly co-regulated with hypoxia and epithelial injury signatures and was more strongly associated with mortality as compared to in the blood. In the healthy human lung, immune tone is closely controlled in order to address the constant inhalation of foreign matter and microbes while also protecting the delicate alveolar architecture necessary for efficient oxygenation.^15–17^ Thus it is not surprising that disruption of pulmonary immune processes has been linked to nearly all pulmonary diseases, including idiopathic pulmonary fibrosis (IPF), chronic obstructive pulmonary disease (COPD), asthma, and hypersensitivity pneumonitis.^11,12^

In contrast, in the blood, the predominant signatures associated with mortality and high-risk lung injury subtypes were those pertaining to endothelial injury, DNA repair, and metabolic signatures of oxidative phosphorylation and energy metabolism. This suggests that the blood compartment may be responding to lung injury by initiating a non-specific hypothalamic-pituitary-adrenal axis program of metabolic support for circulating immune cells, or perhaps showing other general signs of multi-system stress related to extra-pulmonary disease.^18^ Regarding the latter, we note that many patients had poor systemic immunity and comorbidities such as GVHD, colitis, and endotheliopathies that are unfortunately common after HCT. We also note that patients with extreme neutrophilia, as can be seen with acute bacterial infections, was not common in this cohort, likely owing to the clinical practice of prescribing empiric antibiotics rather than using early bronchoscopy, and this may have enhanced the disconnect between systemic and pulmonary inflammation that we observed in our patients.^19–21^

While overlap between BAL and blood pathways of disease were minimal, we did note a few exceptions. A top candidate gene related to post-HCT lung injury in our data was CEACAM6, which was upregulated in both BAL and blood of non-survivors and also tightly co-regulated with BAL expression of numerous genes related to platelet signaling and endoplasmic reticulum stress. CEACAM6 is expressed on the cell surface of respiratory epithelium including airway secretory cells and alveolar epithelial cells,^22^ is upregulated after numerous forms of lung injury, and serves as an intercellular adhesion, anti-apoptotic, and surfactant protective molecule.^23–27^ Future studies addressing the biological role and biomarker utility of CEACAM6 are warranted.

In addition, we did identify a shared BAL-blood transcriptomic signature in lung injury subtype 3, where both the BAL and blood showed upregulation of pathways related to EMT and collagen biosynthesis. Post-HCT fibrosis is a feature of sclerotic GVHD as well as post-HCT bronchiolitis obliterans syndrome and is notoriously difficult to diagnosis in the lungs due to the insidious onset and late imaging findings.^28–30^ Thus our findings could suggest a pathway where detection in peripheral blood might provide insight into pulmonary processes, which could be leveraged for improved diagnostics. It remains unclear whether the detection of this signal in blood represents a synergized systemic fibrotic response or is simply “leakage” of the pulmonary signal into the blood compartment. Future work is needed to identify the most targeted fibrosis biomarkers that might capture both pulmonary and extra-pulmonary disease.

A main finding was the overall lack of transcriptome correlation between paired BAL and blood in this cohort. Supporting this, we identified minimal correlation at the level of individual gene expression, gene set enrichment scores, cell type fractions, and T- and B-cell receptor clonotypes. For example, IL-8 expression in the BAL was not correlated with IL-8 expression in the blood; Hallmark Hypoxia gene set expression in the BAL was not correlated with Hallmark Hypoxia gene set expression in the blood, and the neutrophil fraction in the BAL was not correlated with the neutrophil fraction in the blood. This lack of BAL-blood correlation was true for the overall cohort as well as among subgroups stratified by survival outcome and LIS. Overall, this strongly suggests that transcriptome measurements obtained from peripheral blood cannot be used as a proxy for analogous pulmonary processes and challenges the current clinical paradigm of assessing pulmonary inflammation using blood-based biomarkers.

Other investigators have identified a lack of BAL-blood correlation for cell fractions and cytokines in patients with COVID-19, asthma, and HIV-related lung disease, although transcriptomic comparisons are sparse.^31–37^ Data in HCT patients are rare, although *Kean et al* showed in a non-human primate model of GVHD that peripheral and lung-specific T-cells have notably different TCR repertoires and even different site-specific T-cell transcriptomes when TCRs are shared.^38^ While the lung is well-known to have a robust local immune repertoire, during lung injury, there is degradation of the alveolar-capillary membrane with transmigration of bone-marrow derived immune cells including neutrophils into the alveolar space;^39,40^ thus we were surprised to see the starkly absent cross-site correlation. Since the peripheral blood represents a composite of circulating blood cells as well as signals from all tissues in the body, it is possible that the impact of lung disease on the blood signal was diluted by extra-pulmonary processes. It is also possible that poor post-HCT systemic immune function and/or exogeneous immunosuppression is limiting systemic immune responses to pulmonary processes. Ultimately, the clinical implication of this finding is that peripheral blood testing as a proxy for pulmonary processes may be quite misleading in post-HCT lung injury. Physicians frequently use blood tests of inflammation, such as the C-reactive protein level or the absolute neutrophil count, as a way to infer pulmonary inflammation and guide treatment,^41,42^ but our results suggest this may not be appropriate in the HCT population. Whether more durable markers of pulmonary processes, such as protein biomarkers, can be detected in blood to guide diagnosis and treatment remains an ongoing question in the field.^43–45^

Given these findings, it is likely that the field of post-HCT lung injury will need not only pulmonary-targeted diagnostics, but pulmonary-targeted therapeutics as well. Especially given the significant off-target effects of many immunomodulatory medicines, inhaled therapies coupled with nanoparticles for durable delivery holds high promise for local effect with the least off-target toxicity.^46–48^ However, it remains unclear whether targeting the identified blood-specific pathways associated with disease could actually improve pulmonary outcomes. In support of this possibility, it has recently been shown that targeting intestinal health can improve respiratory health, perhaps through the gut-lung axis.^49,50^

In summary, by comparing 210 paired BAL-blood transcriptomes obtained after pediatric HCT, we identified surprisingly little cross-site correlation in gene expression. Instead, we identified unique site-specific signatures of disease, suggesting compartmentalization of injury-response. These findings strongly support the need for pulmonary-based diagnostics and therapeutics and caution against using peripheral blood testing to guide clinical care in patients with lung injury post-HCT.

## METHODS

### Patients

As previously described,^7^ participating pediatric centers screened all HCT patients preparing to undergo clinically-indicated bronchoscopic BAL for diagnostic assessment of pulmonary disease (NCT02926612). Patients or their guardians were approached prospectively for consent under local IRB approval at each site (UCSF IRB #14-13546, #16-18908) in accordance with the Declaration of Helsinki and permission was obtained to collect leftover BAL fluid as well as paired blood.

### Biospecimen collection

Bronchoscopy and BAL were performed at the discretion of the treating team using local institutional protocols. All BAL were obtained by pediatric pulmonologists trained in fiberoptic bronchoscopy with anesthesia provided by anesthesiologists or critical care physicians. Lavage protocol was not dictated by the study but typically involved 3-6 aliquots of 10mL sterile saline inserted into diseased areas of the lung as determined by preceding chest imaging or physical exam. Percent of lavage returned was not routinely documented and lavage aliquots were typically pooled by the clinical team immediately after collection. After aliquoting for clinical testing, excess lavage remained unfractionated and was placed immediately on dry ice, stored at -70°C until processing. Blood was collected during the BAL procedure, typically within 30 minutes of the lavage; 2.5mL whole blood was collected directly into a PAXgene tube, which was inverted 5-10 times and stored per the manufacturer’s instructions at -70°C until processing.

### Clinical protocols and data collection

Clinical microbiologic testing was determined by the treating team and typically included culture for bacteria, fungus, and AFB; multiplex PCR for respiratory viruses; galactomannan antigen; and cytology for PCP. Additional molecular diagnostics such as PCR for atypical bacteria or fungi were used at the discretion of the site. After BAL, supportive care protocols were determined by the treating team. Patient demographics, medical history, and transplant-specific data were documented by trained study coordinators at each site. The most recent ANC and ALC measured clinically prior to BAL were documented. Results of clinical microbiologic testing on BAL were documented and not considered complete until 4 weeks after collection. Patients were followed until hospital discharge with no loss to follow-up.

### RNA Extraction

BAL underwent RNA extraction as previously described.^8^ Briefly, 200 µL of BAL was combined with 200 µL DNA/RNA Shield (Zymo) and 0.5mm glass bashing beads (Omni) for 5 cycles of 25 seconds bashing at 30Hz, with 60 seconds of rest on ice between each cycle (TissueLyser II, Qiagen). Subsequently, samples were centrifuged for 10 minutes at 4°C and the supernatant was used for column-based RNA extraction with DNase treatment according to the manufacturer’s recommendations (Zymo ZR-Duet DNA/RNA MiniPrep Kit). RNA extraction of PAXgene tubes was performed for this study. Briefly, 1.7mL of peripheral blood mixed with PAXgene reagent underwent 10 minutes centrifugation, washed with PBS, re-centrifuged for 10 minutes, and the pellet was combined with 100uL DNA/RNA Shield, treated with Proteinase K, and underwent a magnetic bead-based extraction with DNase treatment according to the manufacturer’s recommendations (Zymo Quick-RNA Magbead Kit).

### RNA Sequencing

BAL underwent RNA sequencing as previously described using the New England Biolabs Ultra II RNA Library Prep Kit.^62^ First, BAL and peripheral blood RNA underwent sequencing libraries preparation using miniaturized protocols adapted from the New England Biolabs Ultra II RNA Library Prep Kit (dx.doi.org/10.17504/protocols.io.tcaeise). Reagents were dispensed using the Echo 525 (Labcyte) and underwent Ampure-XP bead cleaning on a Beckman Coulter Biomek NX^P^ instrument. The peripheral blood libraries received treatment with rRNA and globin-depleting FastSelect reagent. Libraries underwent 19 cycles of polymerase chain reaction (PCR) amplification, size selection to a target 300 to 700 nucleotides (nt), and were pooled to facilitate approximately even depth of sequencing. Twenty-five picograms (pg) of External RNA Controls Consortium (ERCC) pooled standards were spiked-in to each sample after RNA extraction and before library preparation to serve as internal positive controls (Thermo Fisher Scientific Cat. No 4456740). In addition, to identify contamination in laboratory reagents and the laboratory environment, each batch contained 2 samples of 200 µL sterile water and 6-8 samples of 200 µL HeLa cells taken from a laboratory stock and processed identically to patient samples, in order to account for laboratory- and reagent-introduced contamination. These samples were processed at the same time as the patient BAL samples in order to use the same lot of reagents and minimize batch effect on control samples. Samples were pooled across lanes of an Illumina NovaSeq 6000 or NovaSeq X instrument and sequenced to a target depth of 40 million read-pairs with sequencing read length of 125 nt. Resultant fastq files underwent alignment to hg38 (*STAR*), with mitochondrial, ribosomal, and non-protein-coding transcripts excluded, leading to detection of a median 18,341 protein-coding transcripts per BAL (IQR 17,052-18,755) and a median 14,146 transcripts per blood sample (IQR 13,339-14,560). The dataset was then further subset for samples with >50k total reads to protein-coding genes and genes present in >25% of samples.

### Analysis

Genes differentially expressed by body site were identified by fitting negative binomial generalized linear models to body site with patient grouping as a random effect (*edgeR*).^51^ Genes differentially co-expressed by body site were analyzed at the pathway level by first creating gene set enrichment scores to the MSigDB Hallmark pathways (*gsva*),^52^ calculating geneset-geneset correlation within each site, and then contrasting the correlations across BAL and blood (*DGCA*).^53^ Gene expression correlation across body sites was assessed by subjecting gene counts to variance stabilizing transformation (*DESeq2, vst*)^54^ and then calculating genewise Spearman correlations using paired values. This analysis was repeated using gene set enrichment scores (*gsva*). We then used CIBERSORTx^55^ to impute cell type fractions in BAL using the *Travaglini et al* lung cell atlas^56^ and in blood using the *Schulte-Schrepping et al* blood cell atlas.^57^ We then tested for correlation between BAL and blood cell fractions using Spearman correlations. ImRep^58^ was used to identify T- and B-cell receptor repertoires. Finally, All analyses involving ≥10 comparisons were subjected to FDR adjustment to address multiple hypothesis testing and statistical significance was assessed at the level of FDR-adjusted p<0.05.

Contrasts were then repeated by survival status using each patient’s most recent encounter and by lung injury subtype (LIS). Here, site-specific gene-gene correlation networks were created by identifying genes correlated in one body site (Spearman rho ≥0.5) and not correlated in the other body site (Spearman rho <0.1) with a significant contrast of FDR-adjusted p<0.05 (*DGCA*). Gene networks were plotted using Cytoscape v3.10.3.^59^ Genes involved in each gene network were then ranked by Hub Score (*igraph*)^60^ to identify most central genes, and were summarized at the pathway level using gene set enrichment (*ClusterProfiler*).^61^ We integrated BAL and blood transcriptomes by applying variance-stabilizing transformation (*vst, DESeq2*)^54^ to BAL and blood transcriptomes followed by multi-omics factor analysis (*mofa*),^62^ dimensionality reduction (*umap*), and k-means clustering (*cluster*). Data were visualized using volcano plots (*EnhancedVolcano*), box-whisker violin plots (*ggplot*), heatmaps (*pheatmap*), and gene set enrichment plots (*ClusterProfiler*).

## Data Availability

Raw sequencing files and instructions to request download are available under controlled access on NIH dbGaP: https://www.ncbi.nlm.nih.gov/projects/gap/cgi-bin/study.cgi?study_id=phs001684.v3.p1. Individual-level data are available indefinitely.

## Code Availability

Code and processed data files are available on GitHub: https://github.com/zinterm/pedBMT_BALseq

## ACKNOWLEDGEMENTS

M.S.Z. received research funding from NHLBI K23HL146936, NHLBI R03HL171423, NICHD K12HD000850, the American Thoracic Society, the Pediatric Transplantation and Cell Therapy Foundation, and the National Marrow Donor Program Amy Strelzer Manasevit Grant. M.Y.M received research funding from NCI F31CA271571. H.A-A. received grant funding from the Gateway Foundation and St. Baldrick’s Foundation. J.S.K. and J.J.B. received research funding from NCI P30CA008748. M.A.P. received research funding from NCI P30CA040214. J.L.D. received research funding from the Chan Zuckerberg Biohub. Additional funding for the study was provided by NHLBI UG1HL069254 and a Johnny Crisstopher Children’s Charitable Foundation St. Baldrick’s Consortium Grant.

## AUTHOR CONTRIBUTIONS

M.S.Z., C.C.D., E.M.P., and J.L.D. contributed to the conception and design of the work. All authors contributed to the acquisition of data. M.S.Z., C.C.D., M.Y.M., G.R., M.R.S., E.M.P., H.K., I.S., L.N.S., and J.L.D. contributed to the analysis of data. All authors contributed to the drafting and revision of the manuscript.

