## Supplementary material for "Integrating Pulmonary and Systemic Transcriptomic Profiles to Characterize Lung Injury after Pediatric Hematopoietic Stem Cell Transplant": Table 1 and 2

**Table 1: Patient Characteristics**

| **Patient Characteristics (n=166 patients)** |  |
| --- | --- |
| Age (median years, IQR) | 11.1 (IQR 4.7 - 17.0) |
| Sex (Male) (n, %) | 95 (57.2%) |
| Race (n, %) |  |
| - White | 102 (61.4%) |
| - Black | 20 (12.0%) |
| - Asian/PI | 20 (12.0%) |
| - Other/Multiple | 19 (11.4%) |
| - Native American | 1 (0.6%) |
| - Unknown | 4 (2.4%) |
| Ethnicity - Latino/Hispanic (n, %) | 40 (24.1%) |
| Region (n, %) |  |
| - United States |  |
| - Mid-Atlantic | 18 (10.8%) |
| - Midwest | 24 (14.5%) |
| - Northeast | 9 (5.4%) |
| - South | 11 (6.6%) |
| - Southwest | 11 (6.6%) |
| - West | 67 (40.4%) |
| - Non-US |  |
| - Australia | 21 (12.7%) |
| - Canada | 5 (3.0%) |
| Disease (n, %) |  |
| - Leukemia, Lymphoma | 109 (65.7%) |
| - Inborn errors of immunity | 30 (18.1%) |
| - Non-malignant hematologic | 17 (10.2%) |
| - Solid tumor | 6 (3.6%) |
| - Inborn errors of metabolism | 4 (2.4%) |
| HCT Type (n, %) |  |
| - Allogeneic | 158 (95.2%) |
| - Bone marrow | 69 (43.7%) |
| - Peripheral blood | 63 (39.9%) |
| - Umbilical cord blood (UCB) | 26 (16.5%) |
| - Autologous | 8 (4.8%) |
| HLA match, allogeneic only (n, %) |  |
| - Matched related donor | 31 (19.6%) |
| - Matched unrelated donor (inc. 6/6 UCB) | 37 (23.4%) |
| - Mismatched related donor (haplo) | 44 (27.8%) |
| - Mismatched unrelated donor (inc. <6/6 UCB) | 46 (29.1%) |
| **Event Characteristics (n=210 events with BAL)** |  |
| Days from HCT to BAL (median, IQR) | 131.0 (IQR 37.0 - 375.5) |
| Days from symptoms to BAL (median, IQR) | 8.0 (IQR 2.0 - 22.0) |
| Respiratory support prior to BAL (n, %) |  |
| - No oxygen | 121 (57.6%) |
| - Nasal cannula or Non-invasive positive pressure | 55 (26.2%) |
| - Endotracheal intubation with mechanical ventilation | 34 (16.2%) |
| Comorbidities at time of BAL (n, %) |  |
| - GVHD active at time of BAL | 68 (32.4%) |
| - GVHD ever preceding BAL | 100 (47.6%) |
| - GVHD pulmonary | 20 (9.5%) |
| - Kidney Injury | 32 (15.2%) |
| - Sepsis | 28 (13.3%) |
| - Pericardial Effusion | 16 (7.6%) |
| - Pulmonary Hemorrhage | 15 (7.1%) |
| - Engraftment Syndrome | 13 (6.2%) |
| - TMA | 13 (6.2%) |
| - VOD | 12 (5.7%) |
| - Heart Failure | 9 (4.3%) |
| Immunologic function prior to BAL (median, IQR) |  |
| - ANC (cells/mL) | 3.042 (IQR 1.620 - 5.511) |
| - ALC (cells/mL) | 0.482 (IQR 0.184 - 1.159) |
| Diagnosis without NGS* (n, %) |  |
| - Idiopathic pneumonia syndrome | 111 (52.9%) |
| - Lower respiratory tract infection | 95 (45.2%) |
| - Sepsis | 4 (1.9%) |
| **Outcomes (n=166 patients)** |  |
| Required intensive care (n, %) | 85 (51.2%) |
| Mechanical ventilation >=7 days (n, %) | 48 (28.9%) |
| In-hospital mortality (n, %) | 32 (19.3%) |

**Legend:** Data for each patient (n=166) and each clinical event with BAL and blood collection (n=210) are shown. Count data are described with numbers and percentages; distributions are described with median and interquartile range (IQR).

**Table 2 – Overview of HCT Lung Injury Subtypes**

| **Lung Injury Subtype** | **BAL** | | **Peripheral Blood** | | **Key**  **Insights** |
| --- | --- | --- | --- | --- | --- |
|  | **Gene**  **Expression** | **Gene Co-Expression Networks** | **Gene**  **Expression** | **Gene Co-Expression Networks** |  |
| **Subtype 1**  *(Baseline)* | Alveolar macrophage (AM) predominance, minimal inflammation | N/A | N/A | N/A | Reference subtype with best clinical outcomes |
| **Subtype 2**  *(Pulmonary Neutrophilic Inflammation & Bacterial Infections)* | 2,027 DEGs  increased **granulocyte/ inflammasome** pathways (eg: CXCR1, IL1R2, S100A8)  decreased **macrophage /lymphocyte** signaling  (eg: CCL18, APOE, HLA.DRA) | 9,181 DCEGs  Top hubs: **NLRP3, CXCR1, ALPL**  Pathways: immune activation, cell-cell adhesion, platelet activity, ECM interactions | 1,061 DEGs  increased **thyroid activity** (eg: CALCA, TPO, DIO2)  decreased **heme metabolism** (eg: HBD, HBG2) | 8,347 DCEGs  Top hubs: **NUP210, CDH13, COL6A6**  Pathways: DNA replication, oxidative phosphorylation, cellular metabolism | Pulmonary bacterial infections drive **neutrophil-dominated** lung inflammation and induce a systemic **neurohormonal response** |
| **Subtype 3**  *(Severe Microbiome Depletion & Fibrotic Response)* | 8,166 DEGs  increased **T-cell and fibroblast activation, myogenesis, nitric oxide signaling** (eg: FGF3, BMP1, NOS)  decreased **macrophage & hypoxia signaling** (eg: CD74, FTL, HLA.DRA, HIF1A, HMOX1) | 6,724 DCEGs  Top hubs: **SIGLEC1, PTPRC, HADHB**  Pathways: collagen deposition, integrin interactions, EMT | 574 DEGs  increased **collagen deposition and EMT** (eg: COL4A1, LAMA5, TIMP3). | 9,269 DCEGs  Top hubs: **ANO6, GPD2, TLR8**  Pathways: DNA repair, T-cell signaling, heme metabolism | **Severe microbiome depletion** and **T-cell activation** in lungs coincide with **systemic fibrosis signaling**, suggesting lung injury co-regulates body-wide fibrotic responses |
| **Subtype 4**  *(Viral Infections, Immune Dysregulation & EMT)* | 6,252 DEGs  increased **NK/T-cell activity** **beta-defensins, EMT** (eg: IL2, KLRF1, IFNG, DEFB114, COL11A1, MMP27)  decreased **AM, neutrophil signaling, airway epithelial function** (eg: MARCO, FTH1, MYD88, TREM2, SPRR3, MUC5B, SFTPB) | 10,223 DCEGs  Top hubs: **SLC38A6, ETFDH, TAF2**  Pathways: collagen deposition, ankyrin interactions, EMT | 156 DEGs  weak increased in **IFNα/β expression** (eg: OAS3, IFIT1) | 6,908 DCEGs  Top hubs: **PRR16, SCML2, TRPC**.  Pathways: DNA repair, oxidative phosphorylation, and aerobic respiration | **Severe dysbiosis & viral infections** in the lungs drive **strong pulmonary-specific immune activation and EMT** with a broad systemic metabolic response |

**Legend:** Summary of differential gene expression and co-expression for the post-HCT lung injury subtypes.
